# Reimagining youth-responsive TB care: Voices and visions from adolescents and young adults who completed TB treatment in Nairobi, Kenya

**DOI:** 10.64898/2026.07.02.26357188

**Authors:** Rhoda Pola Karisa, Beate Ringwald, Alice Charity Awinja, Jacqueline Wanjiku Kagima, Drusilla Nyaboke, Justus Simba, Nkirote Mwirigi, Edel Nekesa Sakwa, Stephen Macharia, Aiban Ronoh, Kerry Millington, Immaculate Kathure, Elizabeth Mueni, Chrispine Okoth, Stephen Mulupi, Brenda Mungai, Jeremiah Chakaya, Rachael Thomson

## Abstract

Tuberculosis (TB) services often fail to adequately address the needs of adolescents and young adults in many high-TB-burden settings. Despite evidence on the barriers this age group faces across the TB care pathway, young people are rarely involved in designing solutions to improve care that better meets their needs. This study focused on the perspectives of adolescents and young adults who had completed TB treatment in Nairobi, Kenya, engaging them as partners to examine their treatment experiences and co-create ways to improve youth-responsive TB services.

This participatory qualitative study was conducted among young people aged 15–24 years (n=37) who had completed treatment for drug-susceptible TB from six health facilities in Nairobi County. Other study participants included County TB stakeholders, healthcare workers, facility managers and community representatives (n=214). Data were collected in December 2024 through four participatory workshops organised by age and gender to facilitate open discussion of shared lived experiences. Participants in each workshop used visual methods to describe their journey with TB, identified common challenges through group discussions, and proposed solutions to improve TB services for the young people. The findings from the workshops with the young people were subsequently discussed in stakeholder feedback sessions involving healthcare workers, facility managers, community representatives, and county TB actors. Data were analysed thematically across both workshops and feedback sessions.

Results were structured around four interrelated themes. First, young people reported experiencing prolonged illness before diagnosis, with symptoms often normalised or attributed to other conditions, leading to disruption of school, work, and social life. Second, participants experienced lengthy and uncertain pathways to TB diagnosis, characterised by repeated healthcare visits and treatment for alternative conditions before TB was recognised.

Receiving a TB diagnosis represented a pivotal turning point, evoking both fear and concerns about stigma as well as relief at finally understanding the cause of their illness and accessing treatment. Third, support from family and friends helped young people navigate the challenges of illness, diagnosis, treatment, and repeated health-system encounters. Fourth, participants and stakeholders identified practical service adaptations to strengthen youth-responsiveness, including respectful communication, enhanced counselling, peer support, greater privacy within TB service outlets, and flexible medicine collection such as after 5 pm.

Adolescents and young adults with TB navigate multiple social and health systems barriers before reaching diagnosis, initiating treatment, and completing it. Involving young people directly in identifying gaps and priorities in TB care generated feasible opportunities for strengthening youth-responsive services within existing health systems. Further research is needed to evaluate the effectiveness and acceptability of the models proposed by study participants.

## Introduction

Tuberculosis (TB) remains a major global public health challenge and continues to rank as the leading cause of death from a single infectious agent worldwide [1]. Although the highest burden occurs among adults, adolescents and young adults (AYA) also represent an important but often under-recognised population within TB care and prevention. In 2024, 25% of the global TB notifications were reported from the African region, with approximately 15% (1.8 million) of these occurring among young people aged 15 to 24 years[2]. Despite this burden, adolescents and young adults remain less visible within TB policy, surveillance, and service delivery than young children and older adults[3].

TB prevention and care efforts have historically focused on young children and adults, while the specific needs of adolescents and young adults have received comparatively limited attention [4,5]. This may contribute to delayed diagnosis [6] treatment interruption and poorer treatment outcomes for this age group [7–9]. Recognising these challenges, there have been increasing calls for provision of and stronger attention to adolescent-responsive TB services [10,11]. The World Health Organization updated consolidated TB care and treatment guidelines recommended a youth-centred approach to improve acceptability and continuity of care for young people with TB [11,12].

Adolescents and young adults experience multiple barriers across the TB care pathway. Delayed recognition of symptoms, misconceptions about TB risk, and missed opportunities for early testing can delay diagnosis, while treatment schedules often conflict with work, school, and other social responsibilities[5,13,14]. In some settings, up to 44% of adolescents with TB symptoms do not seek care at health facilities, often due to limited knowledge of TB symptoms, misconceptions, or expectations that symptoms will resolve spontaneously [15]. Social and structural conditions further shape care among young people. Socio-economic factors such as overcrowded living spaces, financial constraints, and stigma may increase vulnerability to TB transmission, progression of TB infection to disease, delays in TB diagnosis, and risk of treatment interruption among young people [14,16,17].

Although these vulnerabilities are increasingly recognised, evidence on how adolescents and young adults experience TB diagnosis and treatment and navigate care remains limited, particularly in low- and middle-income settings where youth-responsive TB services are still evolving. Understanding these experiences can provide practical insights for strengthening person-centred TB care for the population of young people. Few studies have examined the perspectives of young people who successfully complete treatment or explored how their experiences can inform practical service improvements. This study therefore explored the lived experiences of adolescents and young adults who had completed TB treatment in Nairobi, Kenya, and included perspectives from health care workers and county-level stakeholders to identify opportunities for strengthening youth-responsive TB care.

## Methods

This qualitative study formed part of a larger mixed-methods project undertaken in Kenya under the auspices of the LIGHT Consortium research project to understand and address gaps experienced by adolescents and young adults across the TB care cascade. The participatory component reported in this paper centred on the perspectives of AYAs as partners in examining their treatment experiences and co-creating recommendations for more youth-responsive TB services.

The study was conducted between December 2024 and February 2026. Participatory workshops with young people were conducted in December 2024, followed by transcription, data analysis, and interpretation between January and June 2025. Stakeholder feedback sessions were subsequently held at participating facilities between October and December 2025 and culminated in a county-level feedback meeting in February 2026, during which findings and proposed actions were discussed and refined with health system stakeholders.

### Study design

We used a participatory research design that actively involved people with lived TB experience to generate knowledge and information for action. Participatory research recognises that those most affected by a problem are well placed to identify priorities, interpret experiences, and propose contextually relevant solutions [18], including solutions for improving health services, informing program design, and addressing barriers to care [19]. In this study, young people with lived experience of TB used participatory visual methods to share their experiences with TB, including diagnosis, care and treatment, and to identify practical recommendations for strengthening youth-responsive TB services.

Participants discussed their experiences and solutions during feedback sessions with stakeholders, including community representatives, health care workers, sub-county TB managers, and County officials from the Department of Health. Such feedback sessions are increasingly recognised as an important extension of participatory qualitative research because they allow contextual interpretation and co-development of practical action points with health system actors [20].

Participatory approaches were considered particularly appropriate for this study because young people affected by TB often experience stigma, social isolation, and limited opportunities to share their experiences with peers [21,22]. Participatory visual methods facilitated reflection on personal experiences and supported the expression of perspectives that may be difficult to communicate through conventional interview approaches [23]. Bringing young people together in a group setting also created opportunities for peer learning, mutual support, and collective problem-solving, while enabling participants to identify shared challenges and develop recommendations for improving TB services.

#### Study setting

This participatory qualitative study was conducted with adolescents and young adults diagnosed and treated for TB in six health facilities in Nairobi County, Kenya. Nairobi, Kenya’s capital city, has a population of approximately 5.5 million and is one of the largest urban centres in Africa [24]. Although Nairobi’s population is about 10% of Kenya’s population, the city contributes approximately 15% of Kenya’s annual TB burden and records the highest number of TB cases nationally each year [25]. Health service delivery in Nairobi follows Kenya’s tiered health system, with primary care provided through Level II dispensaries and Level III health centres, secondary care through Level IV hospitals, and tertiary referral services through Levels V and VI [26]. The study purposively selected six health facilities whose routine programmatic data showed high TB burden and notable rates of treatment interruption among adolescents and youth. The facilities included: two Level IV public hospitals in Embakasi and Kibra Sub-Counties; two Level III public facilities in Dagoretti and Starehe Sub-Counties; one private facility in Ruaraka Sub-County and one private faith-based facility in Langata Sub-County.

These facilities primarily served populations from informal settlements, characterised by poverty, overcrowding, and limited access to stable income, education, and healthcare services [27].

The research team comprised research, public health, and TB professionals familiar with the Nairobi context. Before data collection, a transect walk with community representatives around selected health facilities was conducted to strengthen contextual understanding of local youth environments and inform interpretation of the findings. [23]

### Study population and participant recruitment

Participants were young people aged 15–24 years who had completed treatment for drug-susceptible TB with a documented treatment completion outcome in facility TB registers. Purposive sampling was used to ensure variation by social context, sex and age group, classified into adolescents (15–19 years) and young adults (20–24 years). The distribution of participants by health facility, age group, and sex is shown in Table 1.

**Table 1.** Distribution of adolescents and young adults participating in the workshops by health facility, age group, and sex (N = 37).

| Health facility | 15–19 Female | 15–19 Male | 20–24 Female | 20–24 Male | Total |
| --- | --- | --- | --- | --- | --- |
| Mama Lucy Hospital | 2 | 2 | 3 | 2 | 9 |
| Mbagathi Hospital | 2 | 1 | 0 | 1 | 4 |
| Riruta Health Centre | 2 | 2 | 0 | 1 | 5 |
| Baraka Health Centre | 2 | 2 | 2 | 1 | 7 |
| St. Mary's Mission Hospital | 1 | 1 | 3 | 0 | 5 |
| Rhodes Chest Clinic | 0 | 3 | 2 | 2 | 7 |
| **Total** | **9** | **11** | **10** | **7** | **37** |

Following identification of eligible participants from facility TB treatment registers by facility health care workers, these persons were contacted by telephone, with support from community health promoters where necessary. During the initial contact, the health care workers provided information about the study and sought permission to share contact details of the potential study participants with the research team. Contact information for interested individuals was then forwarded to the research coordinator, who conducted eligibility screening and provided further information about the study. Individuals who had relocated outside Nairobi or remained unreachable after repeated attempts were considered ineligible.

The social scientist obtained written informed consent from participants aged 18–24 years before participation, after providing an explanation of the study objectives, potential risks and benefits, and participants’ roles and rights to ensure participants fully understood the purpose and nature of the research. For adolescents aged 15–17 years, written consent from a parent or guardian and participant assent were obtained at the health facility before participation. A total of 37 young people were recruited, including 18 young men and 19 young women.

Participants who agreed to participate were invited to workshops at a central location in Nairobi’s business district.

### Participatory workshops

Four participatory workshops were conducted in December 2024 and organised separately by age group (15–19 years and 20–24 years) and by gender (young men and young women) to support open discussions of personal experiences. Each workshop included 9–10 participants in a hotel meeting room, which provided a private and conducive environment for discussions. Workshops were facilitated by the research coordinator (RP), supported by a social scientist (AA) and study administrator (CO). All facilitators had received training in participatory qualitative methods from an experienced social scientist (BR).

We used participatory visual methods to stimulate reflection and discussion among participants. Participants narrated their TB care journey using the River of Life method, a participatory visual method drawing on the analogy of a river and its features [28]. Participants developed individual drawings of a river to represent their life from the onset of symptoms to the present (Fig 1). They represented periods of wellbeing as calm stretches of water with plants or fish, challenges and setbacks as turbulent waters, boulders, or waterfalls, and sources of support as streams joining the main river. Participants then presented their river drawings to the group and reflected on similarities and differences in their experiences.

**Fig 1.**
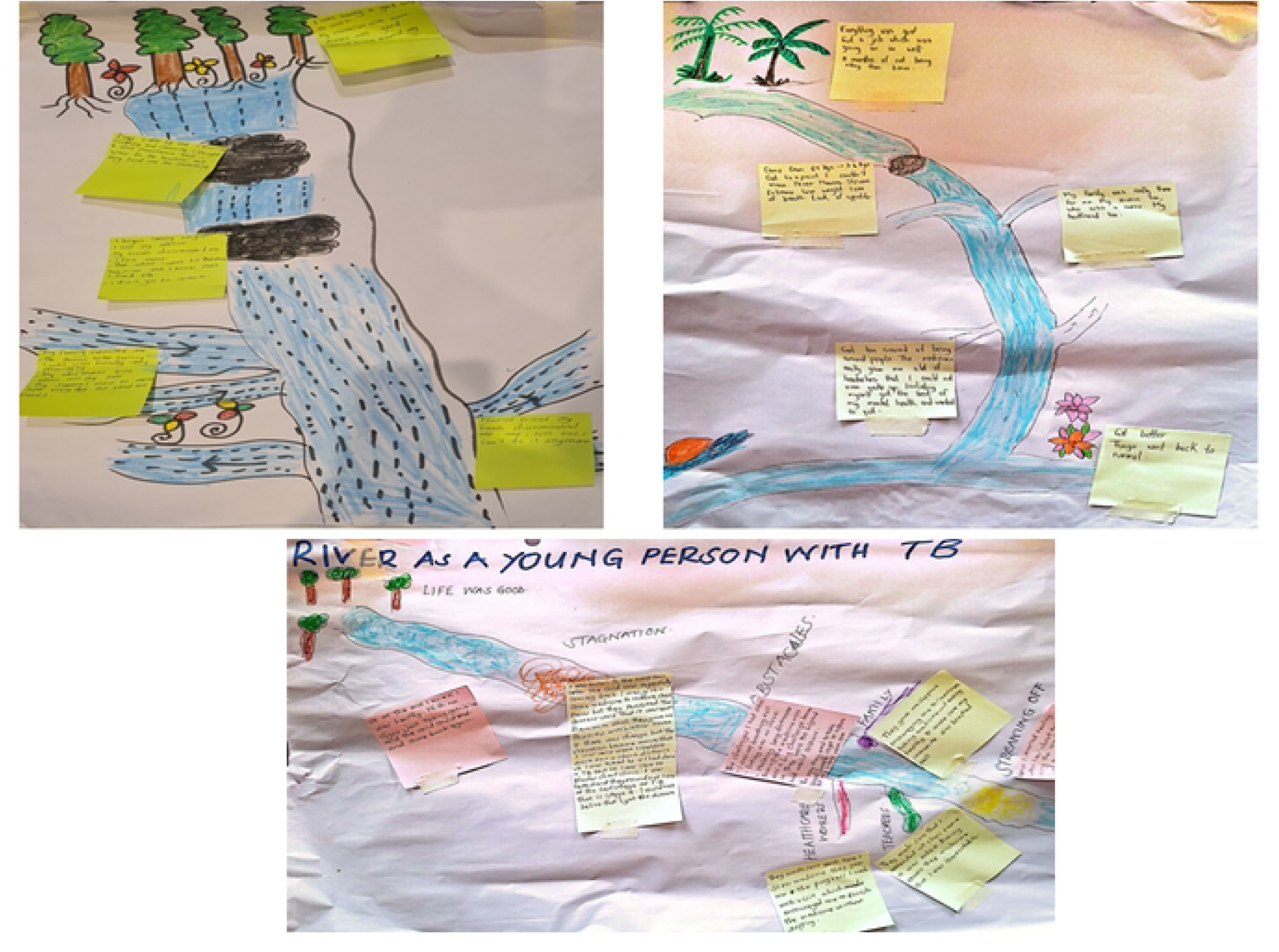
Examples of participant-generated River of Life diagrams illustrating adolescents’ and young adults’ experiences from symptom onset through TB diagnosis, treatment, and recovery. Common challenges identified through the River of Life exercise informed the subsequent Stepping Stones activity. Images reproduced with participants’ consent.

In the second part of each workshop, the Stepping Stones visual tool [23,29] was used to facilitate collective problem-solving. Stepping Stones involved the young people identifying common challenges emerging from the River of Life presentations and working in smaller groups to identify the steps needed to overcome the barriers identified through the River of Life exercise to achieve an outcome (Fig 2). Through facilitated discussion, participants generated practical recommendations for improving youth-responsive TB services.

**Fig 2.**
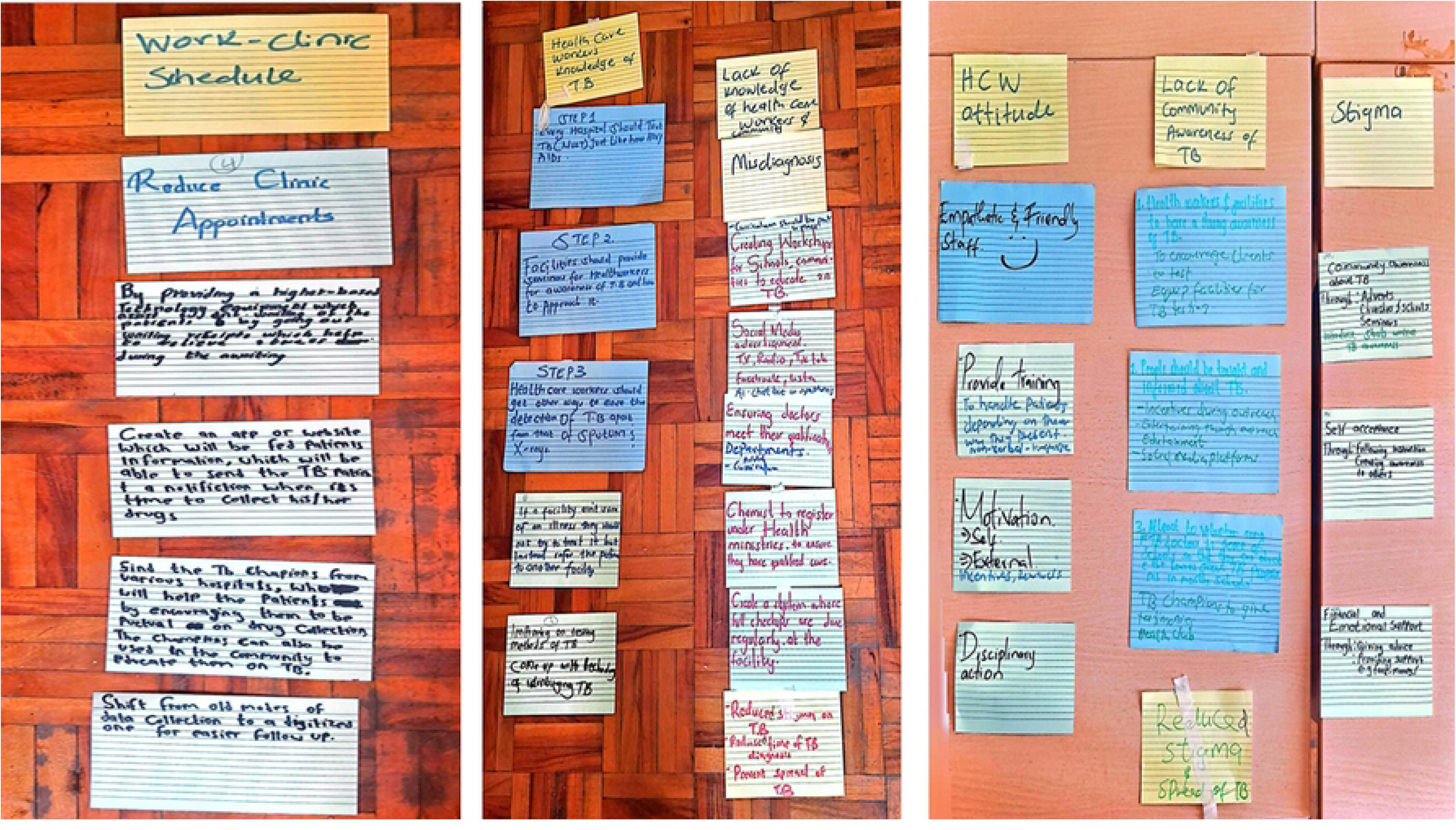
Participant-generated Stepping Stones diagrams illustrating the barriers experienced by adolescents and young adults during the TB care journey and the proposed actions to overcome them. Images reproduced with participants’ consent.

### Stakeholder feedback sessions

Seven stakeholder feedback and action-planning sessions were conducted to share study findings and identify opportunities to strengthen youth-responsive TB services. Six sessions were held at participating health facilities (n=170) and one at the county level (n=44).

During these sessions, the research team and a representative of the study participants presented the thematic findings derived from the River of Life and Stepping Stones activities, including young people’s experiences of navigating TB care, the common challenges they identified, and their proposed solutions. Stakeholders were invited to reflect on the findings, discuss their implications for service delivery, and identify actions that could be implemented to address priorities raised by young people.

Participants in the facility-level sessions included study participants, healthcare workers, facility TB focal persons, sub-county TB coordinators, community health promoters, peer educators, and facility managers. Each facility developed action plans outlining feasible service improvements within their sphere of influence. The county-level session included study participants, TB champions, facility TB focal persons, sub-county health managers, program officers from the NGO sector, and county health managers.

During the county-level feedback session, healthcare workers presented the facility-level action plans developed during the earlier stakeholder meetings, including actions that could be implemented within existing facility resources and those requiring support at sub-county or county level. Participants discussed, refined, and prioritized these actions to inform service improvement efforts.

Participatory workshops lasted approximately six hours, while stakeholder feedback sessions lasted between two and three hours. Discussions were conducted bilingually in English and Swahili to allow participants to express themselves in their preferred language, audio-recorded with participants’ permission, and supplemented by detailed field notes.

### Data management and analysis

Data were analysed using inductive thematic analysis [30]. Analysis was conducted iteratively by members of the research team and involved five stages. First, audio recordings were transcribed verbatim and translated into English prior to analysis by RP and AA. Translations were reviewed by bilingual members of the research team to ensure consistency and preserve meaning. Second, the transcripts, field notes, photographs, and workshop outputs were read repeatedly by RP, AA, and CO to facilitate familiarisation with the data.

Third, initial codes were generated inductively from the data and applied systematically across the transcripts. Coding was conducted independently by RP and AA, with regular discussions with BR to compare interpretations and refine the coding framework.

Fourth, related codes were grouped into broader categories and potential themes. These preliminary themes were reviewed, discussed, and refined by the wider research team to ensure coherence, internal consistency, and alignment with participants’ experiences and perspectives. Finally, themes were defined, named, and organised into a thematic framework that informed interpretation of the findings and development of recommendations.

Stakeholder feedback sessions provided an opportunity for participants and stakeholders to reflect on the study findings, discuss the feasibility of youth-generated recommendations, refine interpretation of the results, and identify priority actions for strengthening youth-responsive TB services. Regular team discussions were held throughout data collection and analysis to reflect on how researchers’ professional backgrounds and assumptions might influence data interpretation.

### Ethical considerations

Ethical approval was obtained from the Kenya Medical Research Institute Scientific and Ethics Review Unit (Protocol No. 4782/KEMRI/RD/22), Liverpool School of Tropical Medicine Research Ethics Committee (23-037), National Commission for Science, Technology and Innovation (NACOSTI/P/24/36765), and the Nairobi County Health Research Committee.

Participation was voluntary. Written informed consent was obtained from all participants, with parental consent and assent for participants below 18 years. Data were anonymised, securely stored, and accessible only to authorised members of the research team. Participants received transport reimbursement of KES 1,000 (approximately USD 7.60).

## Results

Analysis across the participatory datasets identified four interrelated themes reflecting young people’s experiences of TB, the relational and health system factors that shaped their experiences, and priorities for strengthening youth-responsive TB services. These themes draw on insights generated through the River of Life and Stepping Stones exercises and are presented with consideration of similarities and differences by age groups and between young men and young women.

### Living through prolonged illness, uncertainty, and disruption before TB diagnosis

Across all four participatory workshops, the young people illustrated through the River of Life technique that TB is a long, meandering journey often marked by twists, setbacks, and obstacles before a diagnosis was reached. Participants highlighted prolonged periods of uncertainty about the cause of their illness, and disruptions to education, work, and everyday life. Although experiences varied by age and gender, common patterns included delayed recognition of symptoms, sub-optimal self-management of illness, and the social and economic consequences of prolonged ill health.

### Symptoms were prolonged, normalised, and difficult to interpret

Many participants described how symptoms developed gradually and persisted for long periods before TB was suspected. Across all groups, common symptoms included constant cough, chest pain, shortness of breath, fatigue, night sweats, reduced appetite, and weight loss. Several participants initially interpreted symptoms as ordinary respiratory illness or physical exhaustion, particularly when the symptoms temporarily improved. Others experienced unusual manifestations, including inability to walk or persistent vomiting, which were not immediately linked to TB.

> *“My whole body was sick, I could not walk, I was very weak and spent my whole time sleeping, I was unwell for like 2 years.”* (Adolescent, female).
>
> *“I used to get tired every time I worked, coughing and sweating at night, fatigue, night sweats occasionally, shortness of breath.”* (Young adult, male).

Self-medication through chemists, over-the-counter medicines, and home remedies was common, especially during the early stages of illness. For some participants, their health state became alarming only after symptoms worsened significantly, such as coughing blood, severe weakness, and visible weight loss.

> *“I started coughing out a lot of blood, and I got confused and thought my day of death had come.”* (Adolescent, female)

Age and gender differences were evident in how symptoms were interpreted. Adolescents often interpreted the symptoms they faced within the conditions of their school environment, not suspecting serious illness.

> *“Our school was very cold, and we would shower with cold water, so I thought that was the cause of my chest problems.”* (Adolescent, female)

Some young women linked early TB symptoms to common conditions such as colds, tonsils, or pregnancy-related concerns, even when these could be ruled out as causes for poor health.

> “*The symptoms were mistaken for pregnancy, and I was taken by my mother directly for a pregnancy test, which turned negative, and no further action was taken.”* (Young adult, female)

The gradual onset and non-specific nature of symptoms made it difficult for young people to recognise TB as a possible cause of their illness, often delaying decisions to seek further evaluation.

### Illness disrupted school, work, and daily functioning

Persistent symptoms affected young people’s school, college and university attendance, work, and daily functioning. In many Rivers of Life drawings, these disruptions were represented as difficult stretches of the river where progress was slowed by illness, fatigue, and uncertainty. Among adolescents, boys reported that the reduced physical strength limited their participation in sports. Across both genders, repeated absence from school due to ill-health made it difficult to keep up academically.

> *“I was sent home many times from school to be taken to the hospital… but I was not getting better.”* (Adolescent, female)

Older participants more often described interruption of employment, reduced income, and inability to meet their daily responsibilities. Women described missing employment opportunities or struggling to continue working because of physical weakness, while men reported that prolonged illness reduced their ability to work and, in some cases, resulted in job loss.

> *“I missed a job interview because I was too weak to attend, but I was lucky to reschedule and got the job; however, I taught for a few weeks and got worse.”* (Young adult female)
>
> *“My employer did not take my illness seriously. I would faint; my boss did not understand me and would not allow me to go home when I was unwell, and later I was fired.”* (Young adult, male).

In addition to the physical health effects, female participants also felt embarrassed when coughing in class, public transport, or at work, particularly when symptoms drew public attention.

Prolonged illness affected educational participation, employment, and economic stability, while the continued absence of a diagnosis contributed to ongoing uncertainty and disruption in young people’s lives

### Navigating the pathway to TB diagnosis

Across the River of Life drawings, the pathway to a TB diagnosis was commonly represented as a difficult and uncertain stretch of the river marked by waterfalls, boulders, and winding channels that slowed progress towards recovery. Participants described prolonged periods of illness during which they sought care repeatedly, often receiving treatment for other conditions before TB was recognised. For many, obtaining a diagnosis was a significant turning point in the river journey. While the path to diagnosis was characterised by obstacles, uncertainty, and repeated health-system encounters, receiving a diagnosis brought about mixed emotions ranging from fear and concerns about stigma to relief at finally understanding the cause of their illness and gaining access to treatment. These experiences highlight both the challenges young people faced in reaching a diagnosis and the emotional significance of that moment in their TB journey.

### Repeated health-system encounters, financial burdens and system barriers prolonged the pathway to diagnosis

Repeated health system encounters before diagnosis of TB were common across all groups. Participants described prolonged periods of uncertainty during which they sought care from multiple facilities, often receiving treatment for other conditions, including pneumonia, asthma, malaria, and flu, before TB was eventually confirmed. Many reported repeated prescriptions of antibiotics or painkillers without clinical improvement, leaving them frustrated and uncertain about the cause of their illness

> *“After my KCSE [Final high school exam], I went to another hospital. I visited so many hospitals, but in all these places I was given drugs for a cold and pneumonia.”* (Adolescent, female).

Repeated health-system encounters also created financial strain for young people and their families. Multiple consultations, transport costs, diagnostic investigations, and treatment for conditions that were later found not to be the cause of illness placed an additional burden on households already coping with prolonged illness. For some participants, these costs accumulated over months or years before a diagnosis was reached.

> *“My biggest challenge was finances, as a lot of money was used before the diagnosis, while visiting different facilities and treating the wrong disease.”* (Young adult, female)

Participants further described long queues, referral delays, and limited access to diagnostic services as barriers that prolonged the diagnostic pathway. In several cases, young people sought care at higher-level facilities after failing to improve but still encountered delays in obtaining appropriate investigations.

> *“My symptoms were not improving, and that’s where I decided to go to a national hospital, but it was crowded, and I could not wait, so I could not find help.”* (Young adult, male)

Together, these experiences illustrate how repeated healthcare encounters, financial burdens, and health-system constraints delayed access to an accurate TB diagnosis, prolonging both uncertainty and suffering before treatment could begin.

### Receiving a TB Diagnosis brought fear, stigma, and relief

After prolonged uncertainty and repeated healthcare encounters, receiving a TB diagnosis was often described as a major turning point in participants’ Rivers of Life journeys. While the diagnosis explained the persistent symptoms, it also triggered strong emotional reactions. Across all groups, the participants described fear, stigma, disbelief and concerns about death, social relationships and future aspirations. In many of the river drawings, these concerns were represented as branching pathways, reflecting uncertainty about what lay ahead after diagnosis.

> *“I also faced stigma from friends who were afraid to associate with me, with fears that they would get the disease. I had told them about my condition.”* (Young adult, male).
>
> *“I felt like I should die so that the pain could go away; I feared not getting married as a woman, and I lost so much weight.”* (Adolescent female).

For some participants, fears extended beyond TB itself. Young men expressed concerns about HIV and the possibility that others would assume they were HIV-positive because of their TB diagnosis. Others worried about social isolation, rejection by peers, or the long-term consequences of illness on future relationships.

> *“People used to tell me that this could be TB, but I did not want to be tested because I feared I had TB; then I would also have HIV.”* (Young adult, male).

*My wife was also scared that I would infect her, and she started sleeping on the seat. She told me that I have HIV, and she left me.”* (Young adult male).

At the same time, the diagnosis also represented an important point of relief. After months or, in some cases, years of illness, repeated healthcare visits, and unsuccessful treatment for other conditions, confirmation of TB brought clarity and renewed hope. In many River of Life drawings, this moment was represented by calm stretches of water following turbulent sections of the river, symbolising the end of uncertainty and the beginning of a pathway towards recovery.

> *“I was happy when TB was finally diagnosed because I had taken treatment for so many other illnesses with no improvement; I had suffered for about 2 years.”* (Adolescent, female).

Although receiving a TB diagnosis initially generated fear and concerns about stigma, it also explained unresolved symptoms and opened a pathway to treatment, recovery, and renewed optimism for the future.

### Supportive relationships helped young people navigate treatment and recovery

Across the River of Life drawings, family members, friends, and supportive health care workers were often represented as streams feeding into the main river, helping young people navigate the challenges of TB treatment and continue moving towards recovery. Following diagnosis, these supportive relationships provided practical assistance, emotional encouragement, and reassurance during periods of uncertainty. Participants described how family members, peers, and healthcare workers helped them remain engaged in care, adhere to treatment, and maintain hope throughout their recovery journey.

### Family and close relationships supported care seeking and adherence to TB treatment

Family support emerged as a key source of strength throughout the TB journey. Parents were frequently described as recognising worsening symptoms, facilitating healthcare seeking, accompanying participants to facilities, and supporting treatment adherence. Participants who were in school described receiving support from their parents, who would collect the medicines for them and arrange for them to be delivered to school.

> *“I started treatment immediately, and my dad used to collect the drugs for me and bring them to school.”* (Adolescent, female)

This remained important even among older participants living independently, where siblings, partners, cousins, and close friends often became important sources of practical and emotional support. In several accounts, relatives with prior TB experience recognised symptoms early and encouraged further testing.

> “*I got support from my cousin, who told me to go and stay with him; he would cook for me, and my parents kept encouraging me that I would get better.”* (Young adult, male)

These accounts suggest that close relationships played an important role not only in initiating care but also in sustaining treatment through practical and emotional support.

### Supportive TB care strengthened confidence during treatment

Many participants’ experiences with healthcare services became more positive after treatment was initiated. In the River of Life drawings, supportive healthcare workers were often depicted as streams that helped participants move through difficult sections of the river and remain on course towards recovery. Across groups, counselling, follow-up calls, treatment refill flexibility, and respectful interactions with TB clinic staff were viewed as important in sustaining treatment adherence. Counselling was particularly important among adolescent boys and young men who described initial denial following diagnosis before receiving reassurance and encouragement from healthcare workers.

> “*I liked how I could freely talk with the health care workers, and they would call and ask if I am taking my medicines. That really made me feel nice*.” (Adolescent, female)
>
> “*I could pick up my medicines on weekends, and they could give me medicines for a whole week, so that’s how I managed school and picking up medicines. They could give me even excess tabs to ensure that I do not lack medication*.” (Young adult, male)

These experiences suggest that flexible and supportive service delivery may strengthen treatment engagement among adolescents and young adults within routine TB care.

### Young people and stakeholders identified priorities for youth-responsive care

The barriers and challenges represented in the River of Life journeys were linked to the solutions identified through the Stepping Stones exercise. Young people proposed practical actions that could help future adolescents and young adults navigate the pathway to TB diagnosis, treatment, and recovery more effectively. Their priorities focused on improving early diagnosis, making treatment support more responsive to the realities of school, work, and daily life, and reducing stigma through more respectful and confidential care. During stakeholder discussions, healthcare workers, programme staff, and community representatives endorsed these priorities and identified opportunities to strengthen TB services through improvements in provider capacity, more flexible treatment support, and approaches that promote privacy, respect, and peer support for adolescents and young adults.

### Improving early diagnosis and health provider responsiveness

Drawing on their experiences of delayed diagnosis, young people identified limited TB suspicion during routine healthcare encounters as an important barrier to timely care, particularly when symptoms persisted despite repeated treatment. Through the Stepping Stones exercise, participants highlighted several actions that could help improve early diagnosis, including strengthening TB training for frontline healthcare workers, expanding access to diagnostic tools, and promoting earlier referral when symptoms do not improve. During the stakeholder discussions, healthcare workers and TB coordinators supported these priorities and worked with participants to identify feasible actions. Agreed actions included expanding continuing medical education (CME) activities to staff across all departments, providing regular clinical updates on TB, and introducing staff rotations through TB clinics to strengthen practical experience in recognising and managing TB.

> “*It is high time we involve health care workers from all departments in our TB CMEs, not just focusing on the ones in the TB clinic.”* (Sub-County TB coordinator)

These discussions reflected a shared recognition that improving early TB diagnosis requires strengthening provider awareness, diagnostic suspicion, and clinical capacity beyond health care workers in dedicated TB clinics.

### Making treatment support more flexible and youth-friendly

Participants described challenges adhering to TB treatment when clinic schedules conflicted with school, work commitments, and other daily responsibilities. Limited clinic operating hours, particularly during evenings, weekends, and public holidays, often made it difficult to collect medication without disrupting education, employment, or other commitments.

> *“Going for medicine was stressful since I had to move from school in Ngara [Where the school is] to Rhodes [The health facility] and back, yet I felt tired and would sometimes arrive late, and they would be closed*.” (Young adult, female)

Frequent drug collection visits, transport costs, and long waiting times at health facilities further compounded these challenges. Some participants described spending several hours at the clinic, while others reported being asked to return the following day because of incomplete clinical processes, resulting in additional transport costs and missed school or work obligations.

> “*I did not like the long waiting time on drug collection days, or sometimes the health care workers would close and go for lunch*.” (Young adult, male)

In response to these challenges, young people identified steps that would make treatment support more compatible with their daily routines and financial circumstances. Suggested solutions included phone reminders, fewer clinic visits, shorter waiting times, and the use of digital platforms and engagement of champions for follow-up and communication.

Stakeholders supported these priorities and identified practical adaptations that could be implemented within routine services. These included SMS reminders, after-hours or weekend medication collection through departments operating beyond routine clinic hours, and dedicated youth clinic days where adolescents and young adults could access treatment, interact with peers, and receive health education in a more supportive environment.

> “*We will have Fridays as TB clinic day to serve the young people and incorporate some youth talks and health education messages.”* (TB clinic nurse)

Stakeholders also proposed involving community health promoters in home-based or delegated drug delivery for young people who preferred alternative medicine collection arrangements.

> *“We will make use of our Community Health Promoters to deliver drugs for school-going clients at home and at specified locations, as agreed.”* (Community strategy focal person)

Together, these proposed actions reflected a shared commitment to making treatment support for adolescents and young adults should be flexible, convenient, and responsive to their educational, social, and economic realities.

### Reducing stigma through privacy, respectful communication, and peer support

Privacy, confidentiality, and respectful communication were repeatedly identified as important components of positive TB care experiences. Participants described concerns about visible medication packaging, fear of being identified as having TB, stigma within schools and communities, and interactions with healthcare workers that sometimes felt unfriendly or judgmental. These experiences contributed to feelings of embarrassment, discomfort, and reluctance to openly discuss their illness.

> *“I did not like that the drugs I was collecting were not well packed; everyone would see them.”* (Young adult, male)

Young people identified several actions that could help reduce stigma and improve their care experiences. Suggested solutions included discreet medication packaging, more empathetic communication from healthcare workers, and peer-led awareness initiatives in schools, communities, and social media platforms to improve understanding of TB symptoms and reduce misconceptions about the disease.

Stakeholders endorsed these priorities and proposed practical measures to strengthen privacy and youth-friendly communication within routine TB services. These included dispensing medication in plain brown envelopes, engaging youth TB champions in school and community outreach activities, and expanding youth-focused health education through social media and other communication platforms. While stakeholders expressed strong support for these recommendations, they also acknowledged practical resource constraints that could influence implementation. For example, some facilities lacked funds to procure packaging materials and therefore proposed locally feasible alternatives.

> *“Since we as a facility do not have funds to buy envelopes to pack the TB medicines, we will inform our young TB patients to bring their own carrier bags, and we will pack their medicines there.”* (TB clinic nurse)

They also emphasized the importance of respectful provider–patient interactions as a means of building trust and encouraging continued engagement with care.

> *“We will have a youth day at the outpatient clinic where health talks will be provided by a young HCW, and for TB topics, they will include a young TB champion to also talk about their experience.”* (Hospital manager)

These discussions highlighted the importance of creating care environments that protect confidentiality, reduce stigma, and foster supportive relationships between young people and healthcare providers. Both participants and stakeholders viewed these changes as essential for improving the experiences of adolescents and young adults accessing TB services.

Together, the priorities identified through the Stepping Stones discussions demonstrate how adolescents and young adults translated their lived experiences of TB into practical recommendations for improving care. The subsequent stakeholder discussions highlighted opportunities to implement these youth-generated solutions within routine TB services, particularly through strengthening early diagnosis, making treatment support more flexible, and reducing stigma through more respectful and confidential care. These discussions transformed the barriers identified in the River of Life journeys into actionable priorities for strengthening youth-responsive TB services.

## Discussion

This participatory qualitative study highlights that young people with TB in Nairobi experience the disease not only as a biomedical condition but also as a prolonged disruption to education, work, relationships, and everyday social life. Across age and gender groups, delayed diagnosis emerged as a dominant early challenge, often following repeated treatment for other illnesses before TB was recognised. Family members played an important role in navigating diagnosis and sustaining treatment, while young people also identified practical service changes that could make TB care more responsive to their daily realities. By bringing together the perspectives of young people, health care workers, and TB program stakeholders, this study illustrates how individual experiences of TB care and health system responses can interact to shape care pathways for adolescents and young adults, while identifying practical opportunities for strengthening youth-responsive services. The participatory design further enables translation of lived experiences into practical service priorities that were considered feasible within routine care.

A major finding of this study was that the young people often experienced prolonged symptoms before TB was recognised, with most initially interpreting their symptoms to be due to common non-serious respiratory illnesses. Similar findings were reported in a study conducted in Cape Town, where adolescents highlighted downplaying the severity of their symptoms as a contributor to avoidance or delay in care seeking [31]. Many participants initially attempted to manage symptoms using over-the-counter medicines, while others relied on home remedies. Comparable patterns among adolescents and young adults have been documented in other high-burden settings [32,33]. This tendency to normalise symptoms and rely on self-medication may reflect limited TB awareness within households, schools, and the wider community among young people. These findings suggest the need for broader community-based TB awareness strategies that extend beyond health facilities. There is a need to utilise platforms that are accessible to young people, including social media channels such as TikTok and Instagram.

In addition, the prolonged illness led to interference with school, work, and daily activities for the young people, highlighting the broader social consequences of delayed diagnosis. Similar experiences among young people have been described in studies from other high-burden settings, where illness and treatment disrupted school attendance, reduced participation in social activities, and affected the psychosocial well-being [5,22,31,34,35]. These findings underscore the need for TB care models that address not only clinical management but also psychosocial support for adolescents and young adults.

Our findings also show that even after young people sought formal care, diagnosis was often delayed because symptoms were initially attributed to other conditions, resulting in repeated consultations across multiple facilities before TB was confirmed. Similar patterns have been reported in other high-burden settings, where adolescents and children frequently experience multiple provider contacts before a TB diagnosis because TB symptoms overlap with other common illnesses and are not consistently recognised early in the care pathway [31,36].

These delays point to health system gaps in promptly identifying and investigating TB among young people presenting with symptoms compatible with TB. They may also reflect missed opportunities for TB screening across outpatient, emergency, and other departments where adolescents and young adults seek care. Strengthening TB knowledge and clinical suspicion among frontline health service providers across different service points, together with earlier referral and investigation when symptoms persist, may improve timely TB diagnosis for adolescents and young adults.

Repeated visits to multiple health facilities during the care-seeking process imposed significant financial burdens on the young people in this study. Similar findings have been reported in Kenya’s TB patient cost survey, which showed that many TB-affected households continue to experience catastrophic costs despite the availability of free TB diagnosis and treatment services [37]. Evidence from low- and middle-income countries similarly demonstrates that people with TB incur substantial direct and indirect costs while accessing care [35,38–41]. Given the End TB Strategy target of eliminating catastrophic costs for TB-affected households [42], these findings suggest that efforts to strengthen youth-responsive TB care should extend beyond clinical services and include social protection interventions that reduce transport cost, income loss, and other financial barriers experienced by adolescents and young adults and their households [43].

Family support emerged as a central influence across the TB care pathway. Parents, siblings, partners, and other relatives frequently acted as informal care navigators by recognising worsening symptoms, facilitating facility attendance, collecting medicines, and encouraging treatment adherence. Similar findings have been described in TB and chronic disease settings, where family involvement is critical because many adolescents and young people have limited autonomy over health-seeking decisions and treatment logistics [44]. These findings highlight the importance of engaging family members in interventions aimed at improving TB care engagement and treatment outcomes among young people.

Participants described positive experiences with health care workers following TB diagnosis, including counselling, respectful communication, follow-up phone calls and flexible arrangements for drug collection. These forms of support appeared to reduce anxiety and promote continued engagement with treatment. Similar provider practices have been identified as key components of adolescent-responsive care, particularly where young people value respectful communication and continuity and flexibility in service delivery [45–47].

Lessons from HIV programmes further demonstrate that youth-friendly services can enhance engagement and retention in care, highlighting the potential value of adopting such approaches within TB services for adolescents and young adults [48–50].

An important contribution of this study is that young people and stakeholders identified practical service adaptations that could strengthen youth-responsive TB care within existing health systems. Priorities such as flexible medicine collection times and places, youth clinic days, discreet medicine packaging, better provider communication, and peer-led awareness closely align with emerging global recommendations for adolescent-friendly TB services, which emphasise flexibility, privacy, psychosocial support, and care models that minimise disruption to daily life[5,11,35,51].

The adolescents and young people recommended SMS reminders to support adherence amidst competing school, work and social commitments. During the stakeholders’ workshop, this recommendation was prioritised as a feasible action to improve treatment support for the young people. This aligns with evidence suggesting that digital adherence technologies, including SMS reminders, can improve treatment adherence and patient engagement, although findings vary across settings. Given their low cost and scalability, SMS reminders could be integrated into routine TB care to support adherence among adolescents and young adults in resource-constrained settings[52,53].

This study adds context-specific insight into how such interventions may be operationalised within routine facility settings, including the involvement of community health promoters and youth champions to strengthen linkage between facilities and communities. These findings suggest that many youth-responsive adaptations may be achievable within existing service delivery structures when young people are actively involved in identifying feasible solutions. The priorities identified through this study provide practical entry points for strengthening adolescent- and youth-responsive TB services within routine care.

A key strength of this study was the use of participatory research methods, which enabled young people not only to describe challenges but also to generate recommendations for improving TB services. The visual participatory tools, including Rivers of Life and Stepping Stones, supported reflection and expression of experiences that may not have emerged through conventional interviews alone. The subsequent stakeholder feedback sessions provided an opportunity to assess the feasibility of these recommendations and develop context-specific action plans. Together, these processes demonstrate how participatory approaches can bridge the gap between lived experiences and service improvement, generating findings that are both contextually grounded and relevant for policy and practice.

This study has some limitations. First, most adolescent and young adult participants were engaged in care and largely adherent to treatment, which may limit the transferability of the findings to young people who disengage from care or are non-adherent, and whose experiences may differ substantially. Participants who completed treatment may have benefited from more favourable family support, socioeconomic circumstances, or interactions with the health system than those who remained disengaged from care. As a result, the experiences and recommendations identified through the participatory activities may not fully reflect the perspectives and priorities of young people facing the greatest barriers to treatment completion. Second, the study was conducted in an urban setting, and the findings may not be fully transferable to peri-urban or rural contexts, or to counties with different service delivery models and patterns of health-seeking behaviour. Lastly, caregivers were not included in this study, yet they often play an important role in recognising symptoms, supporting care seeking, and influencing treatment adherence among adolescents and young adults. Their perspectives may have provided additional insight into household-level decision-making and support mechanisms across the TB care pathway.

This study highlights that adolescents and young adults experience multiple interconnected challenges across the TB care pathway, from delayed recognition of symptoms and prolonged diagnostic journeys to psychosocial and practical challenges during treatment. At the same time, family support and responsive interactions with health care providers emerged as important enablers of treatment engagement and continuity of care. Through participatory methods, young people and stakeholders identified practical solutions that could strengthen youth-responsive TB services within existing health systems, including greater flexibility, privacy, improved communication, and peer-informed approaches. These findings underscore the importance of embedding adolescent-responsive principles within TB service delivery and demonstrate the value of involving young people directly in the design and improvement of services intended to meet their needs.

## Acknowledgements

We are deeply grateful to the adolescents and young adults who shared their experiences and insights with us. We also acknowledge the support of Nairobi County and the health officials from Langata, Starehe, Embakasi, Kibra, and Dagoretti Sub-Counties, as well as the staff at participating health facilities, for their guidance and collaboration throughout the study.

## Data availability statement

The qualitative data generated and analysed during this study contain potentially identifiable information, including participant-generated visual materials and detailed personal narratives, and therefore cannot be made publicly available in their entirety. Selected de-identified excerpts and participant-generated visual materials reproduced in this manuscript are published with participants’ informed consent. Additional de-identified data supporting the findings may be made available by the corresponding author upon request, in accordance with participants’ consent and institutional data governance procedures.

## Funding statement

This study was funded by the UK Foreign, Commonwealth, and Development Office (Leaving No-one Behind Transforming Gendered Pathways to Health for Tuberculosis). The views expressed do not reflect the UK Government’s official policies. The funders had no role in study design, data collection and analysis, decision to publish, or preparation of the manuscript.

## Competing Interest

The authors declare that they have no competing interests.

## Authors contribution

**Conceptualization of the work**: Rhoda Pola Karisa, Beate Ringwald, Jacqueline Wanjiku Kagima, Drusilla Nyaboke, Stephen Mulupi, Aiban Ronoh, Immaculate Kathure, Stephen Macharia, Jeremiah Chakaya, Kerry Millington, Rachael Thompson, Nkirote Mwirigi

**Methodology:** Rhoda Pola Karisa, Beate Ringwald, Alice Charity Awinja, Drusilla Nyaboke, Stephen Macharia, Aiban Ronoh, Nkirote Mwirigi, Jeremiah Chakaya, Rachael Thompson

**Investigation:** Rhoda Pola Karisa, Beate Ringwald, Alice Charity Awinja, and Chrispine Okoth.

**Data Curation:** Rhoda Pola Karisa, Beate Ringwald, Alice Charity Awinja, Chrispine Okoth

**Formal analysis and interpretation of data:** Rhoda Pola Karisa, Beate Ringwald, Alice Charity Awinja, Chrispine Okoth, Elizabeth Mueni, Brenda Mungai

**Funding Acquisition:** Stephen Mulupi, Kerry Millington, Jeremiah Chakaya, Chrispine Okoth, Rachael Thompson

**Writing original draft:** Rhoda Pola Karisa

**Review and revision of the article:** Rhoda Pola Karisa, Beate Ringwald, Alice Charity Awinja, Jacqueline Wanjiku Kagima, Drusilla Nyaboke, Justus Simba, Edel Nekesa Sakwa, Stephen Mulupi, Aiban Ronoh, Immaculate Kathure, Elizabeth Mueni, Chrispine Okoth, Stephen Macharia, Brenda Mungai, Jeremiah Chakaya, Kerry Millington, Rachael Thompson, Nkirote Mwirigi

**Supervision and oversight**: Aiban Ronoh, Stephen Macharia, Immaculate Kathure, Drusilla Nyaboke, Chrispine Okoth, Jeremiah Chakaya, Elizabeth Mueni, Beate Ringwald, Rachael Thompson

## Notes

### Author Declarations

Ethical approval was obtained from the Kenya Medical Research Institute Scientific and Ethics Review Unit (Protocol No. 4782/KEMRI/RD/22), Liverpool School of Tropical Medicine Research Ethics Committee (23-037), National Commission for Science, Technology and Innovation (NACOSTI/P/24/36765), and the Nairobi County Health Research Committee. Participation was voluntary. Written informed consent was obtained from all participants, with parental consent and assent for participants below 18 years. Data were anonymised, securely stored, and accessible only to authorised members of the research team

